# Effects of vitamin D supplementation and seasonality on circulating cytokines in adolescents: analysis of data from a feasibility trial in Mongolia

**DOI:** 10.1101/19001842

**Authors:** Sergey Yegorov, Sabri Bromage, Ninjin Boldbaatar, Davaasambuu Ganmaa

## Abstract

Vitamin D deficiency is prevalent in human populations and has been linked to immune dysfunction. Here we explored the effects of cholecalciferol supplementation on circulating cytokines in severely vitamin D deficient (blood 25(OH)D_3_ << 30 nmol/L) adolescents aged 12-15 from Mongolia. The study included 28 children receiving 800 IU daily cholecalciferol for 6 months spanning winter and spring, and 30 children receiving placebo during the same period. The levels of 25(OH)D_3_ were assessed at baseline, three and six months. Twenty-one cytokines were measured in serum at baseline and at six months. The median blood 25(OH)D_3_ concentration at baseline was 13.7 nmol/L (IQR=10.0-21.7). Supplementation tripled blood 25(OH)D_3_ levels (p<0.001) and reversed the direction of change for most cytokines (16/21, 86%). Supplementation was associated with elevated interleukin (IL)-6 (p=0.043). The placebo group had reduced MIP-1α (p=0.007) and IL-8 (p=0.034) at six months. These findings suggest that cholecalciferol supplementation and seasonality have a measurable impact on circulating cytokines in adolescents, identifying chemokines as potentially important biomarkers of vitamin D status in this population.

**ClinicalTrial.org ID:** NCT01244204

## INTRODUCTION

Accumulating evidence indicates that vitamin D has important non-skeletal functions, particularly in the immune system (1-5). In particular, vitamin D deficiency has been associated with increased risk for diseases tightly linked to immune function, such as autoimmune conditions and respiratory tract infections (1, 4-6). Notably, a recent meta-analysis found that vitamin D supplementation significantly reduced the risk of acute respiratory infections most prominently in individuals with low blood 25(OH)D concentrations (<25 nmol/L) (7), and in our earlier studies in Mongolian children, in whom vitamin D deficiency (25(OH)D_3_ <25 nmol/L or 10 ng/ml (8)) prevalence exceeds 80%, we observed a significant reduction of acute respiratory tract infection associated with cholecalciferol supplementation (9, 10). Similar observations were made regarding the relationship of vitamin D and immunity to TB infection in other cohorts (2).

The *in vivo* effects of vitamin D on immunity are incompletely understood in the context of different human populations and genetic backgrounds (3), although studies performed *in vitro* and in animal models indicate that the nature of vitamin D effects on immunity is context- and cell type-dependent. For example, vitamin D exerts stimulatory effects on monocytes and macrophages inducing interleukin (IL)-1 production, while modulating adaptive immune responses by increasing IL-10 production by dendritic and T cells (1, 5).

Given the protective effects of vitamin D against respiratory infections and the evidence of the vitamin’s immune roles *in vitro*, here we tested the hypothesis that six months of cholecalciferol supplementation in vitamin D-deficient adolescents would result in a significant change in the circulating mediators of antiviral and antibacterial immunity.

## METHODS

### Study setting and participant recruitment

This study is part of a feasibility pilot trial (completed in 2010) to assess the effects of vitamin D supplementation on latent TB incidence in 120 Mongolian children (ClinicalTrial.org ID: NCT01244204) (9). The screening and recruitment of participants was described previously (9, 11). Briefly, children aged 12-15 years residing in Ulaanbaatar, the capital of Mongolia, were recruited. The study intervention consisted of 800 IU vitamin D or placebo (Tishcon Corp., Salisbury, MD) daily for 6 months from November 2009 to May 2010, during the coldest 6 months of the Mongolian year. For the current study 58 paired serum samples (baseline and six months) were randomly selected from the parent placebo (n=30/59) and supplemented (n=28/61) groups. This study was designed to supply pilot data for future immune studies in the same cohort, therefore no formal sample size calculations were performed and sample size was determined based on the available study budget. All study procedures were approved by the institutional review boards of the Mongolian Ministry of Health, National University of Mongolia and the Harvard School of Public Health (Ref. #16571). Written consent to participate was collected from both children and their parents.

### Sample collection and diagnostic testing

Blood (8.0 ml) was collected by venipuncture into red top tubes (Becton Dickinson). Serum was isolated by centrifugation and stored at -80^0^C prior to shipping for analysis to the United States. Measurement of 25(OH)D_3_ was performed using Liaison 25(OH)D_3_ Vitamin D total assay at 0, three and six months. The concentrations of 21 cytokines representing chemokines (macrophage inflammatory protein (MIP)-1α, MIP-1β, MIP-3α, interleukin (IL)-8, fractalkine, interferon-inducible T-cell alpha chemoattractant (ITAC), homeostatic (IL-2, IL-7, granulocyte-macrophage colony-stimulating factor (GM-CSF), classic proinflammatory (IL-1β, IL-6, tumour necrosis factor (TNF), Th17-type proinflammatory (IL-17α, IL-23), regulatory (IL-10, IL-21) as well as Type I (interferon (IFN)γ, IL-12) and Type II cytokines (IL-13, IL-4, IL-5) (see **Supplementary Table 1** for details) were measured at baseline and at 6 months using the Human High Sensitivity T Cell Panel (HSTCMAG-28SK) on a MAGPIX instrument (EMD Millipore). All experimental assays were performed by research personnel blinded to the supplementation status of participants.

### Statistical analysis

Differences in demographic characteristics between the cholecalciferol supplemented and placebo groups were assessed using Independent-Samples Mann-Whitney U and Chi-Square Tests. Mean height-for-age and height-for-BMI z scores were calculated using WHO Anthro software (12). Cytokine concentrations were Log10-transformed prior to analysis to facilitate visualization, while hypothesis testing was performed on the original (not transformed) data. Serum 25(OH)D and cytokine concentrations across study visits were compared using paired t-test, while differences between the placebo and cholecalciferol supplemented groups at each time point were assessed by one sample t-test. To compare the cumulative change in cytokine concentrations between the supplemented and placebo groups, we first calculated the proportion of cytokines that were found on average increased or decreased at follow-up for each participant group (solid dots for each cytokine in **Figure 2**), and then performed a Chi-Square test of the null hypothesis that there was no significant change in cumulative cytokine concentrations between the placebo and cholecalciferol-supplemented children. All statistical analyses and graphing were performed using IBM SPSS V.23 (NY, US) and GraphPad Prism V.6.0. (CA, US).

## RESULTS

### Participant demographics

Samples from a total of 58 children were analyzed. At baseline the median serum 25(OH)D_3_ concentration was 13.7 nmol/L and basic socio-demographic characteristics were not significantly different between the supplemented and placebo groups (**Supplementary Table 2**).

### Changes in systemic 25(OH)D_3_ concentrations

First, we examined blood 25(OH)D_3_ levels at baseline (November) and three and six months (February and May, respectively) (**Figure 1**). Children receiving placebo had significantly reduced 25(OH)D_3_ in February (mean= 10.6 nmol/L, mean fold change (MFC)=1.4, p<0.001) and an increase of 25(OH)D_3_ in May (mean= 23.2 nmol/L, MFC=1.5, p<0.001) compared to November. On the other hand, children receiving cholecalciferol exhibited an elevation of 25(OH)D_3_ in February (mean= 44.3 nmol/L, MFC=3.1, p<0.001) and this increase was sustained in May (mean= 49.0 nmol/L, MFC=3.5, p<0.001) compared to November. Compared to the placebo group, cholecalciferol supplemented participants had significantly higher blood 25(OH)D_3_ levels in both February (4.2-fold, p<0.001) and May (2.1-fold, p<0.001) (**Figure 1**).

**Figure 1.**
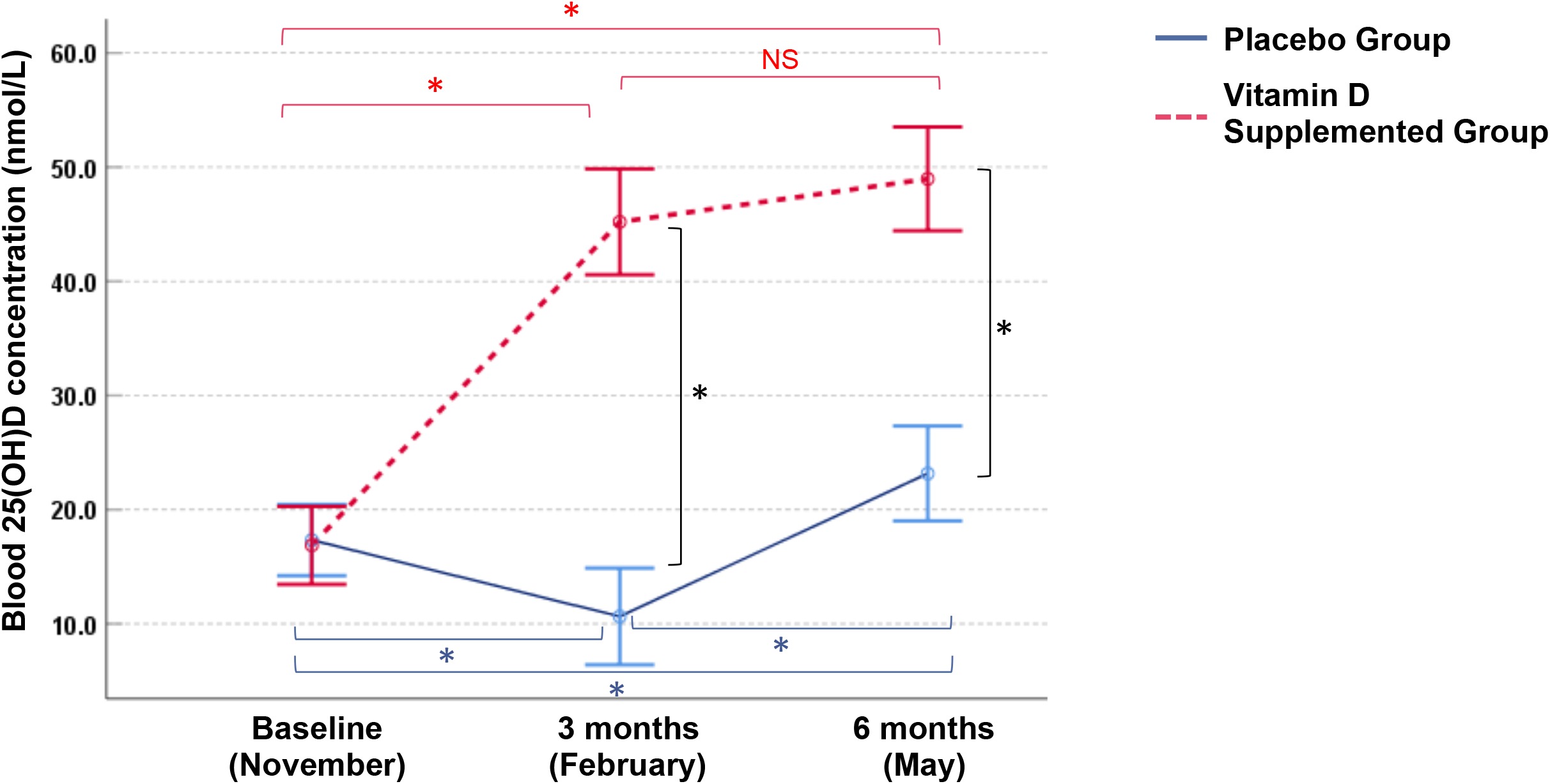
Longitudinal changes in blood 25(OH)D concentrations in Mongolian children. Measurements were performed in vitamin D3 supplemented (n=25) and placebo (n=30) groups. Circles and bars denote means and 95% confidence intervals, respectively. Intraindividual changes within each group and inter-group differences were assessed by paired t test and one-sample t test (p< 0.05), respectively. *p values <0.001, NS=not significant.

**Figure 2.**
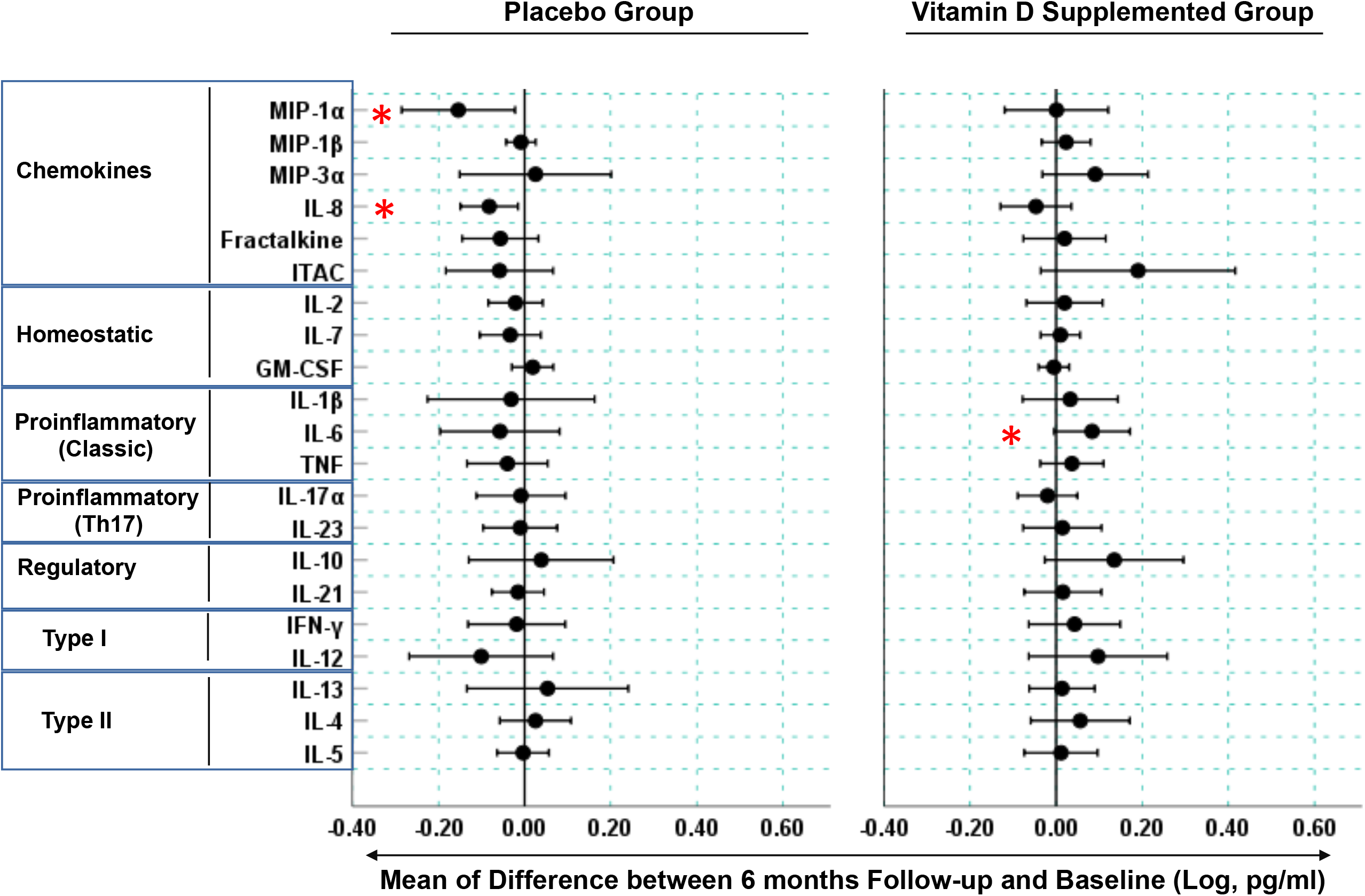
Longitudinal changes in blood cytokine concentrations in Mongolian children. Measurements were performed on serum samples from vitamin D3 supplemented (n=22) and placebo (n=27) groups. Circles represent means of difference between Log-transformed cytokine concentrations at the six-month follow-up visit and baseline. Bars are 95% confidence intervals. Cytokine change is considered significant when its respective confidence intervals are found entirely on the positive or negative sides of the x-axis and are not spanning the “0” reference line. Stars denote cytokines (MIP-1α, IL-8, IL-6) exhibiting statistically significant changes (p< 0.05)- see **Supplementary Figure 1** for participant-level data for these cytokines.

### Changes in systemic cytokine concentrations

Due to missing data, paired cytokine analysis was performed for 49 (27 placebo and 22 supplemented) participants. First, we compared intra-participant changes in cytokine concentrations between baseline and six months after study initiation (**Figure 2**). In the supplemented group, 85.7% (18/21) and 14.3% (3/21) of measured cytokines exhibited a positive and negative change, respectively, while in the placebo group the mean difference in cytokine concentrations was negative for 76.2 % (16/21) and positive for 23.8% (5/21) of the assessed cytokines. Thus, cholecalciferol supplementation had a significant effect on the cumulative change in blood cytokine concentrations compared to placebo (Chi-square statistic= 16.2, p<0.001). The cholecalciferol supplemented group had elevated IL-6 (MFC=1.4, p=0.043) (**Figure 2** and **Supplementary Figure 1**), while the levels of chemokines MIP-1α and IL-8 were significantly reduced in the placebo group (MFC= 2.38, p=0.007 and MFC=1.31, p=0.034 for MIP-1α and IL-8, respectively, **Figure 2** and **Supplementary Figure 1**). The mean cytokine concentrations did not significantly differ between the placebo and supplemented groups at baseline or at six months.

## DISCUSSION

In this study the supplemented group’s 25(OH)D_3_ concentrations increased gradually reaching 49 nmol/L at six months, while the un-supplemented adolescents remained severely deficient with blood 25(OH)D_3_ levels remaining <25 nmol/L throughout the study, linking the observed cytokine changes with the recent meta-analysis findings indicating that 25 nmol/L is a critical threshold of vitamin D deficiency associated with elevated risk for acute respiratory infection (7). Interestingly, in May the placebo group had higher 25(OH)D_3_ concentrations compared to November, although these levels were still under the critical threshold of 25 nmol/L (**Figure 1**)

Compared to baseline, six months of cholecalciferol supplementation resulted in a cumulative shift toward elevation for a majority (∼%86) of measured cytokines. This was in stark contrast with the effect seen in the placebo group, where compared to baseline ∼76% of cytokines exhibited a cumulative shift toward reduction, suggesting that vitamin D deficiency could contribute to a gradual change of immune variables over winter. Further, the vitamin D-deficient children had significantly reduced MIP-1α (CCL3) and IL-8 (CXCL8) concentrations at six months compared to baseline. This effect was not seen in the supplemented participants, suggesting that vitamin D deficiency is implicated in the reduction of these chemokines playing roles in leukocyte homing and activation and mediating antibacterial and antiviral immune responses. Previously, vitamin D supplementation in the context of chronic kidney disease was implicated in decreasing the concentrations of another chemokine, monocyte chemoattractant protein-1, and our findings provide further evidence for the role of vitamin D in chemokine homeostasis (13).

The increased IL-6 observed in the supplemented children is consistent with *in vitro* studies showing that macrophages and monocytes up-regulate IL-6 expression upon vitamin D stimulus (1) and IL-6 expression is up-regulated in the peripheral blood mononuclear cells of vitamin D supplemented multiple sclerosis patients (14). At the same time vitamin D supplementation was found to down-regulate IL-6 in some studies (15-18). These seemingly contradictory findings regarding IL-6 and vitamin D could be attributed to various confounders and differences among the studies, such as the duration and dosage of vitamin D supplementation, the participants’ genetic background, underlying clinical conditions and extent of vitamin D insufficiency/deficiency at baseline.

Our findings are consistent with other studies reporting an increase of IL-10 in vitamin D supplemented individuals (19-21), as we also observed a strong trend to elevated IL-10 in the supplemented children. Somewhat surprisingly, we saw no change in IFN-γ in our study, the Type I cytokine that was significantly elevated in vitamin D-supplemented US and Mexican adults (19, 20), which could reflect the diverse effects of vitamin D in different age and/or ethnic groups.

Our findings should be interpreted in the light of limitations. First, the study was designed as a pilot with a small sample size, which must have reduced the power to detect small differences in cytokine concentrations. The small sample size also limited our choices for data analysis, and we were unable to employ multivariable modeling in this study. Our analysis was also limited by the cytokine panel size and time points included in the study. Lastly, we tested the effects of a rather conservative vitamin D regimen and future studies should assess the effects of different supplementation protocols on systemic immune mediators.

In summary, this study for the first time assessed the immune effects of cholecalciferol supplementation in adolescents from North/East Asia. Although it is challenging to differentiate between the effects of vitamin D supplementation versus seasonality, six months of cholecalciferol supplementation had a perceivable impact on systemic immune mediators in this population. Future studies should explore in more detail the utility of the identified cytokines as biomarkers of vitamin D-mediated immune function and the associations of the identified immune pathways with clinical outcomes.

## Data Availability

All data generated or analyzed during this study are included in this published article.

## DECLARATIONS

## Acknowledgements

This study was supported by the NIH/NHLBI 1K99HL089710-01A1 grant to GD. We acknowledge the support of Harvard Catalyst - Laboratory for Innovative Translational Technologies (HC-LITT) at Harvard Institutes of Medicine with cytokine assays. We thank all the participants and research teams involved in the study. We are especially grateful to Yaruuna Enkhbold, Sonom Boldbataar, Jeremy Furtado, Allison Halleck, Gansuvd Balgansuren, Purevdorj Olkhanuud and Winston Patrick Kuo for their technical advice and expertise. We thank Dr. Adrian Martineau for the comments on the earlier draft of the manuscript.

## Availability of data and materials

All data generated or analyzed during this study are included in this published article.

## Authors’ contributions

Conceptualization, Sergey Yegorov and Davaasambuu Ganmaa; Data curation, Davaasambuu Ganmaa; Formal analysis, Sergey Yegorov and Sabri Bromage; Funding acquisition, Davaasambuu Ganmaa; Investigation, Sergey Yegorov, Sabri Bromage, Ninjin Boldbaatar and Davaasambuu Ganmaa; Methodology, Ninjin Boldbaatar; Project administration, Davaasambuu Ganmaa; Resources, Davaasambuu Ganmaa; Supervision, Davaasambuu Ganmaa; Visualization, Sergey Yegorov; Writing – original draft, Sergey Yegorov; Writing – review & editing, Sabri Bromage, Ninjin Boldbaatar and Davaasambuu Ganmaa.

## Ethics approval and consent to participate

All study procedures were approved by Harvard School of Public Health IRB and Mongolian Ministry of Health ERB. Written informed consent was obtained from all children and their parents.

## Supplementary information

Supplementary information for this article includes Supplementary Tables 1 and 2 and Supplementary Figure 1.

